# Prevalence of untreated prediabetes and glucose metabolism disturbances in Mexico: An analysis of nationally representative surveys spanning 2016-2021

**DOI:** 10.1101/2023.05.16.23290081

**Authors:** Carlos A. Fermín-Martínez, César Daniel Paz-Cabrera, Martín Roberto Basile-Alvarez, Paulina Sánchez Castro, Alejandra Núñez-Luna, Jerónimo Perezalonso-Espinosa, Daniel Ramírez-García, Neftali Eduardo Antonio-Villa, Arsenio Vargas-Vázquez, Luisa Fernández-Chirino, Karime Berenice Carrillo-Herrera, Leslie Alitzel Cabrera-Quintana, Rosalba Rojas-Martínez, Jacqueline A. Seiglie, Omar Yaxmehen Bello-Chavolla

## Abstract

**OBJECTIVE:** Characterizing prediabetes phenotypes may be useful in guiding diabetes prevention efforts; however, heterogeneous criteria to define prediabetes have led to inconsistent prevalence estimates, particularly in low- and middle-income countries. We estimated trends in untreated prediabetes prevalence in Mexico across different definitions and its association with prevalent cardiometabolic conditions.

**METHODS:** We conducted a serial cross-sectional analysis of National Health and Nutrition Surveys in Mexico (2016-2021), totaling 21,442 Mexican adults, excluding individuals with diagnosed or undiagnosed diabetes. Prediabetes was defined using ADA (impaired fasting glucose [IFG] 100-125 mg/dL and/or HbA1c 5.7-6.4%), WHO (IFG 110-125 mg/dL), and IEC criteria (HbA1c 6.0-6.4%). Prevalence trends of prediabetes over time were evaluated using Poisson regression and its association with prevalent cardiometabolic conditions with logistic regression.

**RESULTS:** Prevalence of prediabetes in Mexico in 2021 was 27.3%. Despite an overall downward trend in prediabetes (RR 0.960, 95%CI 0.940-0.979), this decrease was primarily driven by decreases in prediabetes by ADA-IFG (RR 0.883, 95%CI 0.861-0.907) and WHO-IFG criteria (RR 0.878, 95%CI 0.838-0.921), while prediabetes by ADA-HbA1c (RR 1.049, 95%CI 1.022-1.077) and IEC-HbA1C criteria (RR 1.064, 95%CI 1.014-1.115) increased over time. Prediabetes prevalence increased over time in adults >40 years, individuals with central obesity, self-identified as indigenous or living in urban areas. Regardless of the definition, prediabetes was associated with increased risk of cardiometabolic conditions.

**CONCLUSIONS:** Prediabetes rates in Mexico from 2016-2021 varied based on defining criteria but consistently increased for HbA1c-based definitions and high-risk subgroups. Regardless of the definition, prediabetes clusters subjects with high cardiometabolic risk.

## INTRODUCTION

Prediabetes is a stage of altered glucose metabolism associated with risk of progression to diabetes mellitus and an elevated burden of cardiovascular risk factors (1,2). While comprehensive data on the global burden of prediabetes are lacking, the International Diabetes Federation estimates a global prevalence of Impaired Glucose Tolerance (IGT) of 7.5% (374 million adults), of whom 72.2% reside in low- and middle-income countries (LMICs). Ethnicity is a known modifier of metabolic risk in individuals with prediabetes (3,4); in particular, Mexican mestizo populations have an increased risk of diabetes, which manifests at younger ages and lower body-mass index levels (5–7). However, epidemiological data on prediabetes among ethnically diverse populations are limited, including in Mexico, where diabetes mellitus is a leading cause of disability and death (8). Given that prediabetes is the earliest identifiable stage of glucose dysregulation and progression to diabetes mellitus can be prevented at this stage (2,9,10), a more nuanced understanding of prediabetes epidemiology in Mexico could help inform diabetes prevention efforts (2).

Despite being a potentially reversible risk factor for diabetes, an important challenge in the timely identification of prediabetes in clinical and public health contexts lies in the heterogeneity and controversy of its definition (2,11,12). Five definitions of prediabetes have been proposed by professional societies and are in current practice (13–15). The American Diabetes Association (ADA) defines prediabetes as either fasting plasma glucose (FPG, 100-125 mg/dL), glycated hemoglobin A1c (HbA1c, 5.7-6.4%), or two-hour post 75-g oral glucose load (140-199 mg/dL)(15). The World Health Organization (WHO) and the International Expert Committee (IEC) proposed definitions that include two-hour post 75-g oral glucose load or FPG (WHO) or HbA1c-only (IEC) with distinct cut-offs (13,14). These discrepancies have led to heterogeneous estimates of prediabetes prevalence and conflicting data on the utility of its identification (2,12,16). In this study, we aimed to provide reliable estimates on prevalence trends of untreated prediabetes in Mexican adults from 2016 to 2021 using a series of nationally representative surveys, and to assess potential modifiers of prediabetes prevalence. Finally, we also quantified the extent to which prediabetes is associated with prevalent cardiometabolic comorbidities.

## METHODS

### Study design

We conducted a serial cross-sectional analysis using data from the Mexican National Health and Nutrition Survey (ENSANUT) for the years 2016, 2018, 2020, and 2021 (17–20). Briefly, ENSANUT is a population-based survey that aims to assess the health and nutritional status of Mexican adults. ENSANUT is representative at a national, regional, and rural/urban level, as it uses two-stage probabilistic cluster stratified sampling based on households and individuals. Participants underwent a comprehensive questionnaire collecting demographic, socioeconomic, and health-related data, and a physical exam including measurement of blood pressure and anthropometry. A random subsample of each cycle had an additional biochemical evaluation with serum samples for fasting glucose, insulin, lipid profile, and glycated hemoglobin (HbA1c); these biomarkers were used to estimate prevalence of glucose metabolism disturbances. For this study, ENSANUT 2006 and 2012 were not included because of insufficient biomarkers to perform adequate classification of alterations in glucose metabolism. Details on study design and a complete flowchart for the selection of participants are outlined in **Supplementary Materials**.

### Variable definitions

#### Diagnosed and undiagnosed diabetes

Previously diagnosed diabetes was defined by self-report among individuals who answered “yes” to the question “Has a doctor ever told you that you have diabetes (or high blood sugar)?” or report of using insulin or oral diabetes medications. Individuals who did not meet the criteria for prior diagnosis, but who had either fasting plasma glucose (FPG) ≥126 mg/dL or hemoglobin A1c (HbA1c) ≥6.5% were categorized as having undiagnosed diabetes. The prevalence of diabetes according to these definitions was estimated using individual-level data from all ENSANUT cycles.

#### Prediabetes and insulin resistance

We defined prediabetes using two different biomarker-based definitions. Impaired fasting glucose (IFG) was defined using fasting plasma glucose levels between 100-125 mg/dL and high HbA1c was defined using HbA1c levels between 5.7-6.4%, as proposed by the ADA (21). To further evaluate concordance and discordance of these criteria, we defined variables for individuals with IFG but HbA1c within the normal range (IFG-Normal A1c), individuals with elevated HbA1c but normal fasting glucose (NFG-High A1c), and individuals who fulfilled both criteria (IFG-High A1c). Furthermore, we defined insulin resistance (IR) as HOMA2-IR values ≥2.5, as previously defined for the Mexican population (22,23). Finally, we sought to study the impact of higher diagnostic thresholds by using the prediabetes definitions proposed by the WHO (FPG 110-125 mg/dL) and IEC (HbA1c 6.0-6.4%) (13,14). For all these definitions, we excluded individuals with previously diagnosed or undiagnosed diabetes mellitus. Prediabetes definitions used for this study are summarized in **Table 1**.

**TABLE 1.**
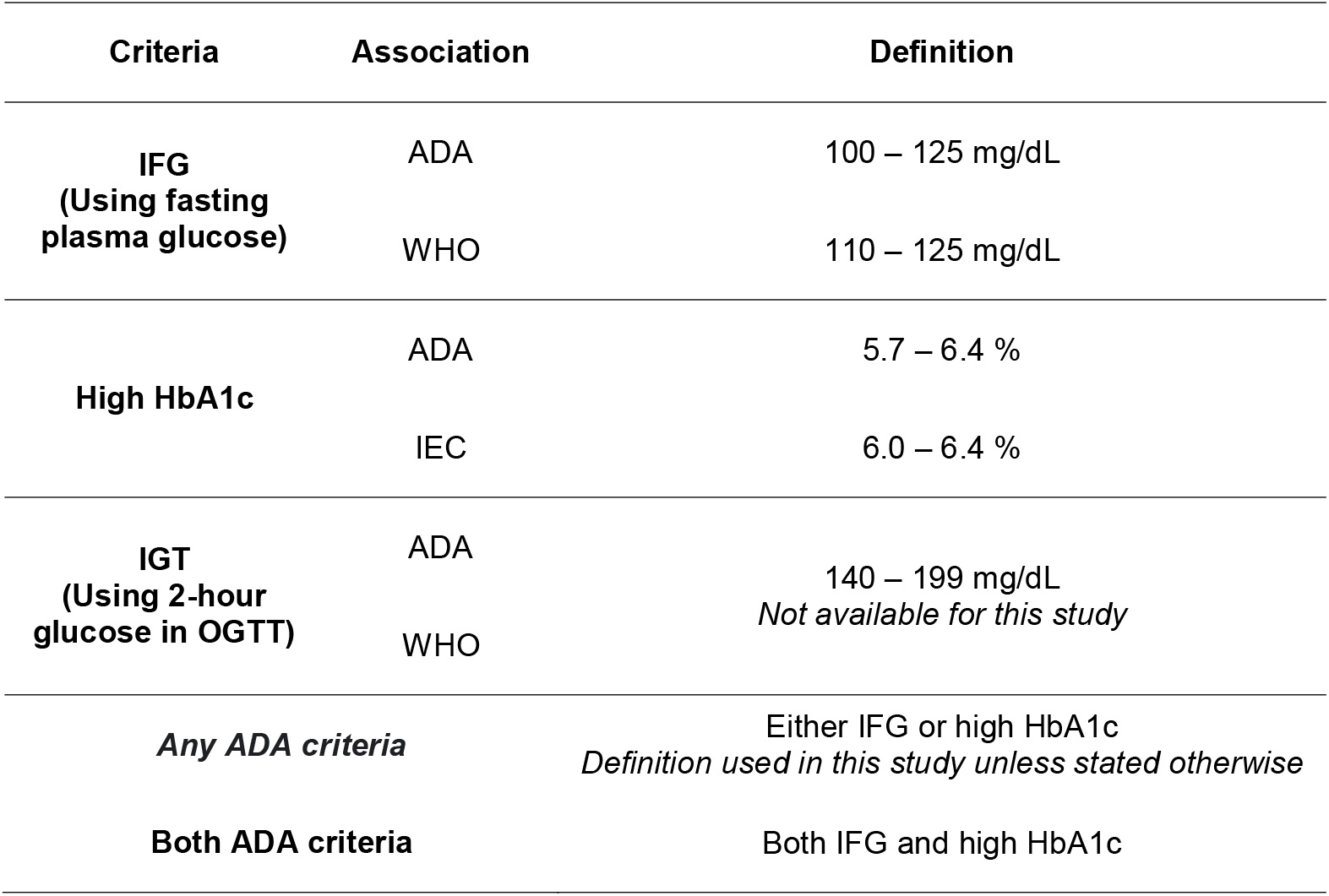
Prediabetes as defined by the ADA, WHO and IEC using various diagnostic tests. The default definition used throughout this study —unless stated otherwise— is having either IFG or high HbA1c as defined by the ADA. <u>Abbreviations</u>. ADA: American Diabetes Association. WHO: World Health Organization. IEC: International Expert Committee. IFG: Impaired fasting glucose. IGT: Impaired glucose tolerance. OGTT: Oral glucose tolerance test.

#### Covariates

We used body mass index (BMI) to categorize patients with normal weight (18.5-24.9 kg/m^2^), overweight (≥25 kg/m^2^) or obesity (≥30 kg/m^2^). We also used waist circumference M≥90 cm for men or ≥80 cm for women to define central obesity based on IDF criteria (24).

To assess state-level social disadvantage we used the 2020 density-independent social lag index (DISLI) by obtaining the residuals from a linear regression of population density onto social lag index, which is a composite measure of access to education, health care, dwelling quality, and basic services in Mexico (25–27).

#### Cardiometabolic conditions

Hypertension was defined by either a self-reported prior diagnosis, use of blood pressure-lowering medications, a systolic blood pressure ≥140 mmHg, or a diastolic blood pressure ≥90 mmHg (using the mean of at least two different measurements). Hypercholesterolemia and hypertriglyceridemia were defined by fasting total cholesterol ≥200 mg/dL or fasting triglycerides ≥150 mg/dL, respectively. Metabolic syndrome was defined using IDF criteria: obesity (by either BMI or waist circumference) plus two of the following: triglycerides ≥150 mg/dL (or specific treatment for this abnormality), HDL-cholesterol <40 mg/dL for men or <50 mg/dL for women (or specific treatment), arterial blood pressure ≥130/85 mmHg (or specific treatment), FPG ≥100 mg/dL or prior diagnosis of type 2 diabetes (24). Cardiovascular disease was defined as a self-reported prior diagnosis of either myocardial infarction, heart failure or stroke.

### Statistical analyses

#### Weighted prevalence of prediabetes and its modifying factors

Prevalence of prediabetes and insulin resistance were estimated using sample weights from ENSANUT for participants with available HbA1c measures; all estimations were conducted using the *survey* R package (28). We further performed weighted subgroup analyses for prevalence trends stratified by age category (20-39, 40-59 or ≥60 years old), sex, BMI category, central obesity, smoking status (never-smoker, former-smoker, or current-smoker), indigenous identity, rural or urban area, and DISLI category (high or low/middle). To determine whether these modifying factors had an influence on prediabetes trends over time, we implemented mixed-effects Poisson models where the interaction term between ENSANUT year and each factor was included as a fixed effect and the state of origin as a random intercept to address state-level conglomeration. We used population estimates by state from the National Population Council (CONAPO) (29), which we included as an offset to model annual rates of prediabetes and to obtain prevalence rate ratios.

#### Prediabetes as a predictor of cardiometabolic conditions

To assess whether participants with prediabetes (using the ADA, WHO and IEC definitions) showed an increased risk of cardiometabolic conditions compared to euglycemic individuals, we fitted logistic regression models for all ENSANUT years adjusting for age, sex, and BMI. The outcomes for these models were arterial hypertension, hypercholesterolemia, hypertriglyceridemia, insulin resistance, metabolic syndrome, and cardiovascular disease. We did not exclude participants with diabetes to compare the risk conferred by this condition with that of prediabetes. The only exceptions were the models for insulin resistance and metabolic syndrome, as the presence of diabetes substantially influences these definitions. All statistical analyses were conducted using R version 4.1.2 and p-values thresholds are estimated for a two-sided significance level of α = 0.05.

## RESULTS

### Study population

We included adults aged ≥20 years old who completed the health questionnaire, totaling 66,641 participants (2016: 8,301, 2018: 42,565, 2020: 2,373, 2021: 13,402). Among them, 21,442 were selected for venous blood sampling (2016: 3,913, 2018: 13,004, 2020: 2,373, 2021: 2,152). A flowchart diagram of participant selection is available in **Supplementary Figure 1**. We observed a consistent predominance of women (57.5 %) and an overall median age of 44 years (32–57 years). The proportion of participants with indigenous identity ranged from 4.7 % (in 2021) to 11.6 % (in 2016) and those living in urban areas ranged from 50.1 % to 76.4 %. We also observed a slight, but significant, increment in BMI and waist circumference throughout the years. A complete outline of population characteristics is shown in **Supplementary Table 1**, and percentages of missing values for each variable are depicted within **Supplementary Figure 2**.

### Trends in the prevalence of glucose metabolism disturbances in Mexico

Prevalence of prediabetes by any definition was higher than that of diabetes in Mexico between 2016 and 2021. However, the pattern we observed was irregular, with a steep decrease from 2016 to 2018 (26.3% vs. 19.0%), followed by a marked increase from 2020 to 2021 (19.2% vs. 27.3%, **Figure 1A**). Poisson models showed an overall decline in prediabetes prevalence per year (RR 0.960, 95%CI 0.940-0.979); however, we observed an increase using ADA-HbA1c criteria (RR 1.049, 95%CI 1.022-1.077) and a decrease with ADA-IFG criteria (RR 0.883, 95%CI 0.861-0.907). A similar overall trend was observed for the stricter WHO-IFG (RR 0.878, 95%CI 0.838-0.921) and IEC-HbA1c criteria (RR 1.064, 95%CI 1.014-1.115), indicating that the overall decreasing trend for prediabetes using any criteria was driven by decreases in IFG which offset increases in high HbA1c. To further characterize this phenomenon, we classified individuals with prediabetes into those who presented with both high HbA1c and IFG and those who displayed only one of these alterations (**Figure 1B**). Among participants with discordance in these criteria, we observed an increase in the proportion of high HbA1c and a decline in IFG over the years, with a consistent increase in individuals with concordant IFG-high HbA1c, likely indicating increases in sustained alterations in glucose metabolism (**Figure 1B**). Furthermore, individuals with prediabetes with alterations in both parameters had the highest proportions of insulin resistance (IR), comparable to the IR levels of subjects with diabetes (**Figure 1C**).

**FIGURE 1.**
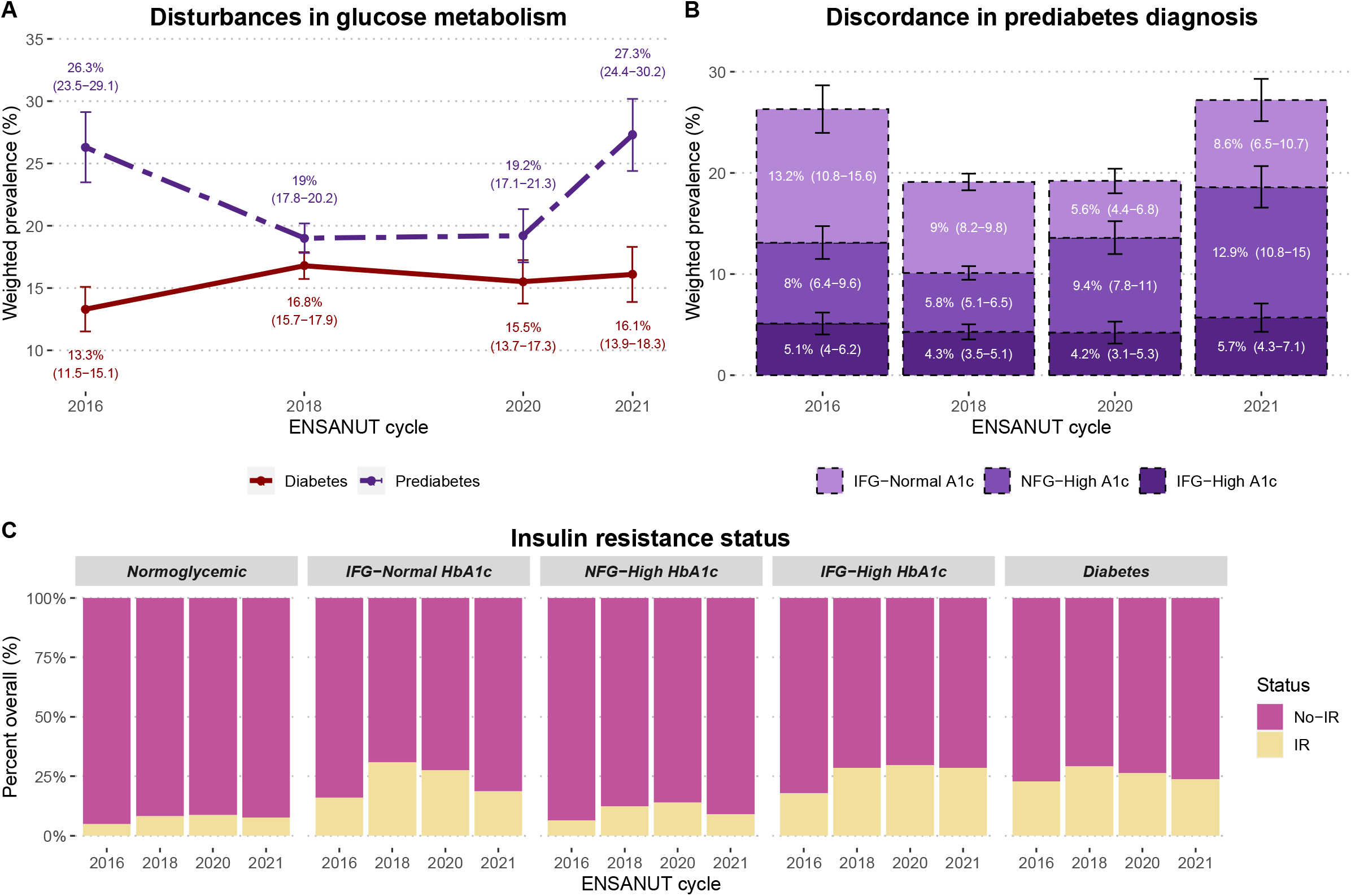
Change in prevalence of diabetes and prediabetes from 2016 to 2021 in Mexico (A). Participants with prediabetes (either high HbA1c or IFG) are categorized according to the diagnostic criteria they fulfilled (IFG only, high HbA1c only, or both), revealing that changes in prevalence were primarily driven by subjects with high HbA1c (B). The percentage of participants with insulin resistance (HOMA-2IR ≥2.5) in each category is also shown (C). <u>Abbreviations</u>. IFG: Impaired fasting glucose. NFG: Normal fasting glucose. IR: Insulin resistance.

### Determinants of prediabetes prevalence in Mexico

Several factors significantly increased the risk of prediabetes such as older age, BMI ≥25 and ≥30 kg/m2, and central obesity. Current smokers had a lower risk than never smokers, but this could be attributed to reverse causality. Moreover, although the overall tendency was a decrease, some groups displayed an increment in prediabetes prevalence over the years, namely participants ≥40 years, those with central obesity, and those living in urban areas (**Figure 2, Table 2**). When assessing only high HbA1c as the diagnostic criteria for prediabetes, we found that indigenous identity and being a former smoker (compared to never smokers) were additional risk factors (**Supplementary figure 3, Supplementary tables 2-3**). Lastly, to address potential explanations for the observed differences in prediabetes prevalence related to racial and ethnic background, we contrasted rates of insulin resistance and various demographic and health-related parameters between participants with and without indigenous identity. Briefly, we observed lower rates of insulin resistance among indigenous subjects; however, results had greater heterogeneity in this population. Notably, indigenous participants displayed higher age, DISLI, HbA1c and triglycerides, Indigenous participants also displayed overall lower HOMA2-IR and HOMA2-β indicating, on average, lower β-cell function (**Supplementary figure 4**).

**FIGURE 2.**
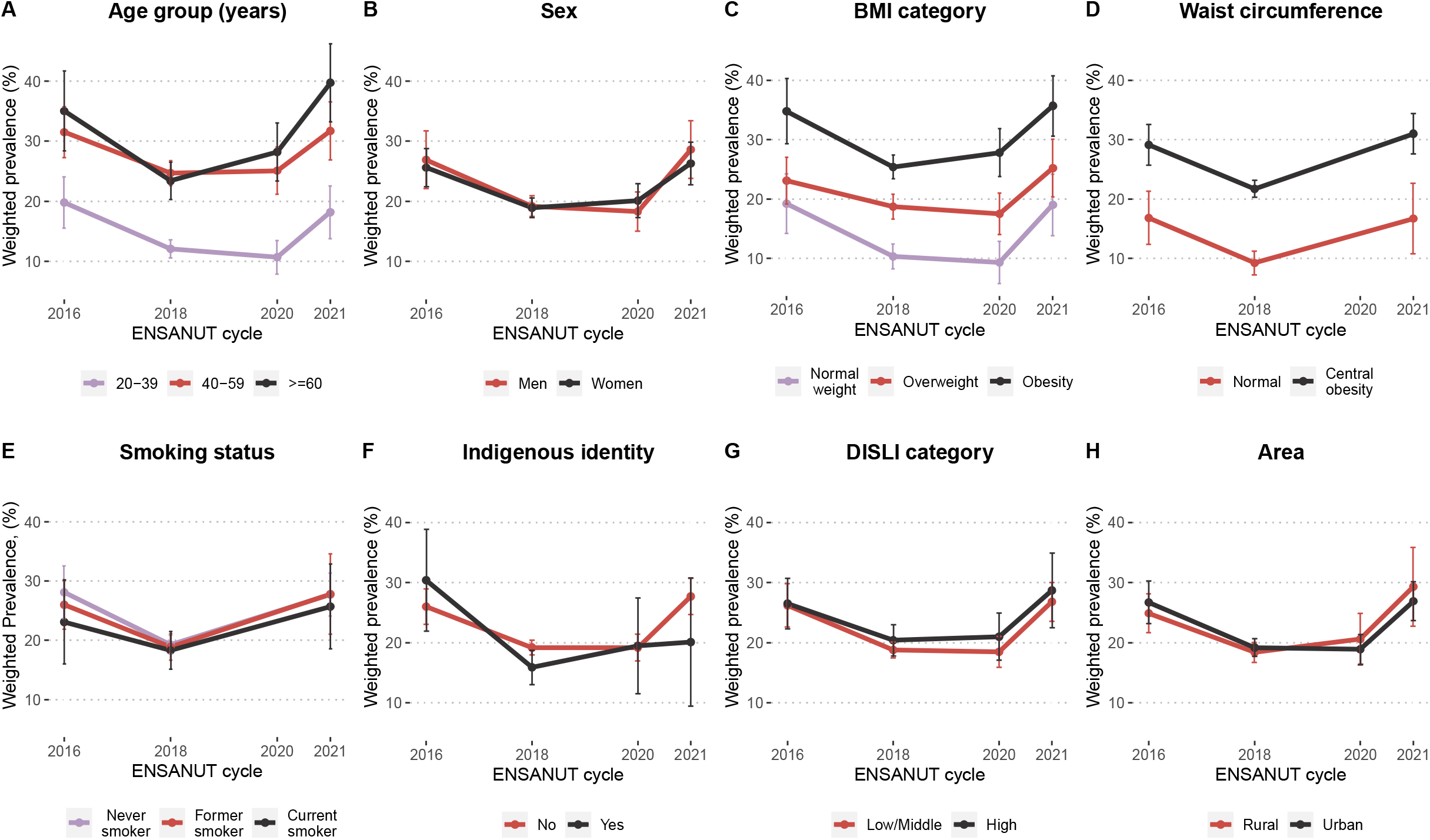
Change in prevalence of prediabetes (either high HbA1c or IFG) from 2016 to 2021 in Mexico stratified by age group (A), sex (B), BMI category (C), waist circumference (D), smoking status (E), indigenous identity (F), DISLI category (G) and area (H). <u>Abbreviations</u>. BMI: Body mass index. DISLI: Density-independent social lag index. IFG: Impaired fasting glucose.

**TABLE 2.**
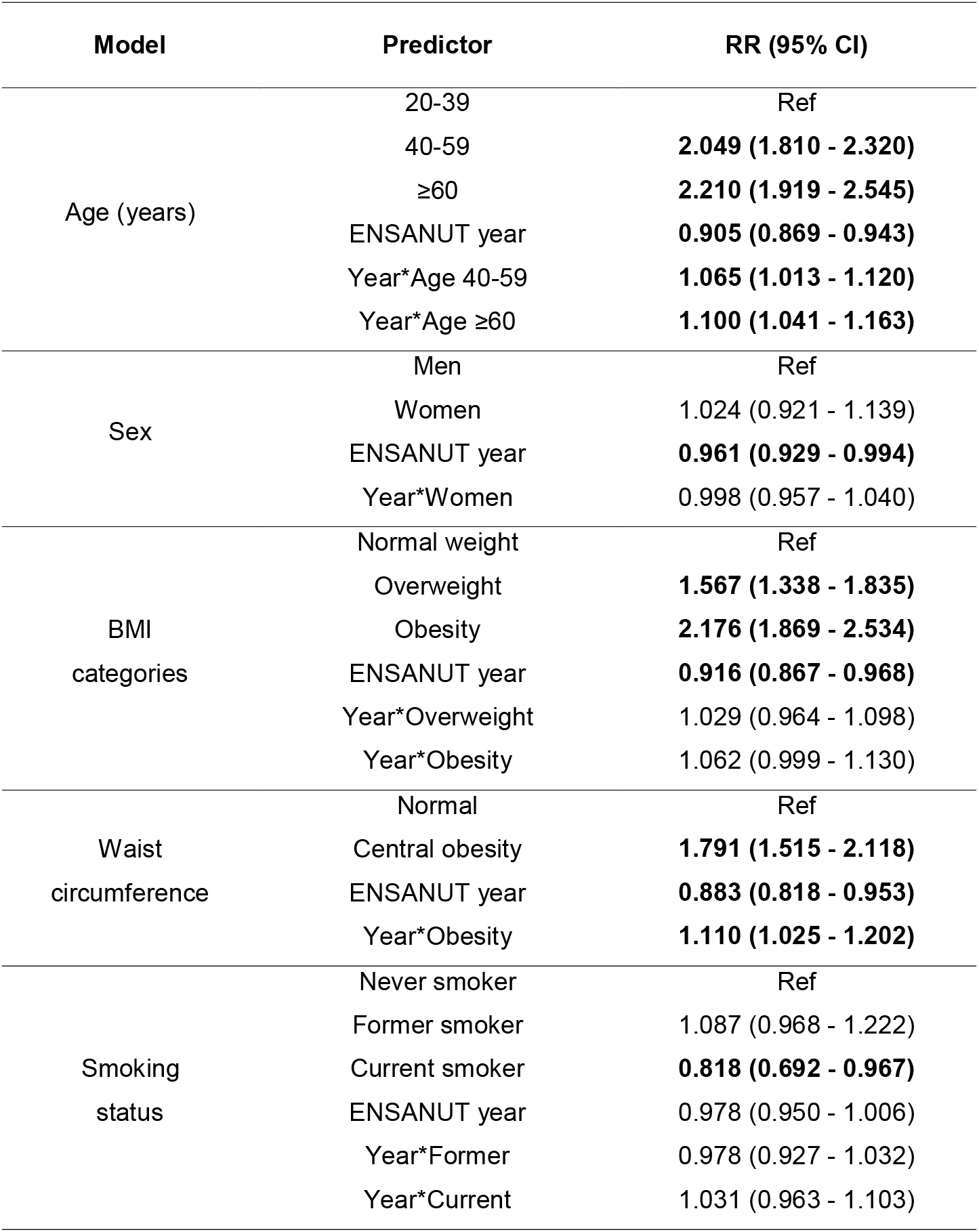
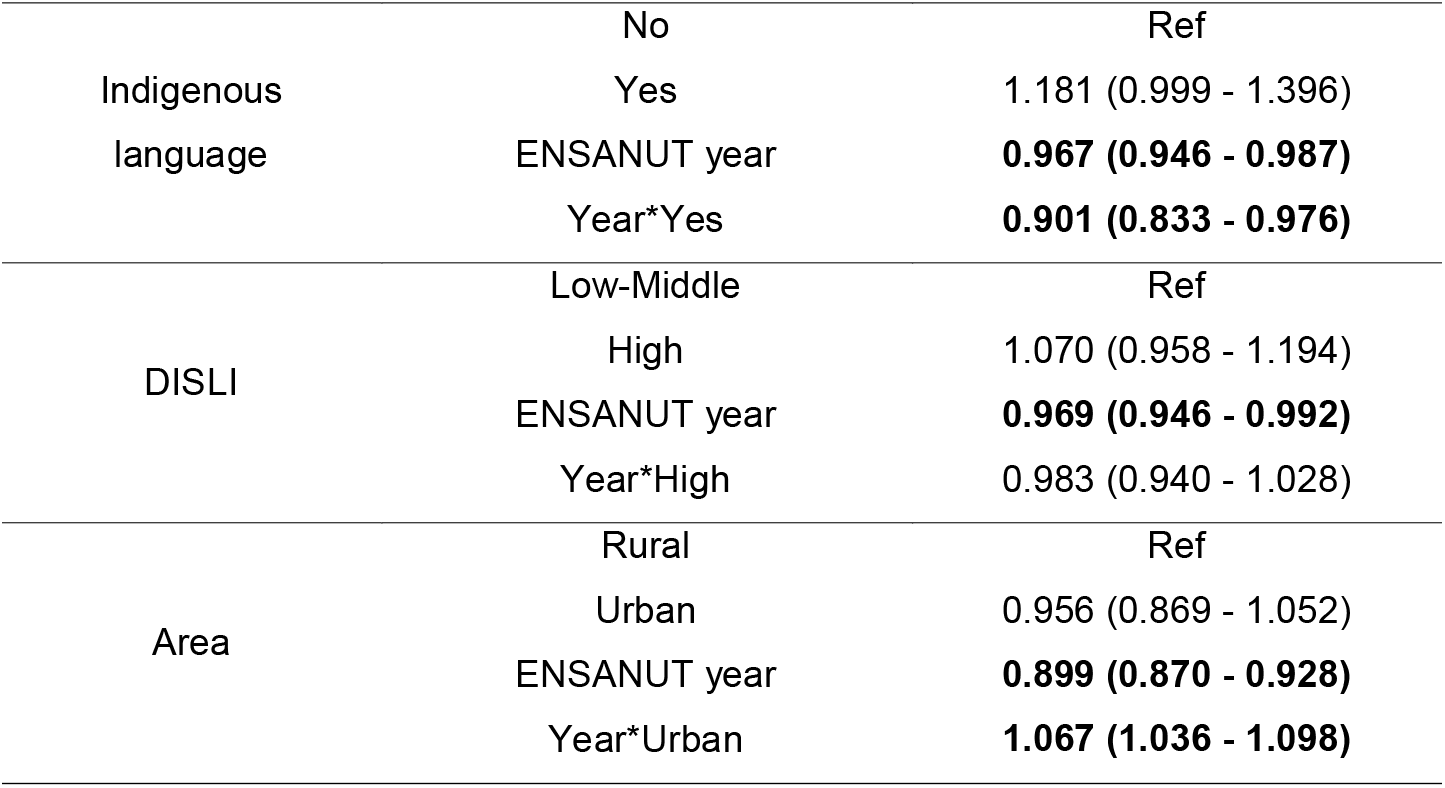
Prevalence rate ratios (RR) with 95% confidence intervals from mixed-effects Poisson models to quantify the influence of several modifiers on prediabetes rates over time (either high HbA1c or IFG). <u>Abbreviations</u>. BMI: Body mass index. DISLI: Density-independent social lag index.

### Prediabetes as a predictor of cardiometabolic conditions

We estimated the prevalent cardiometabolic risk (age, sex and BMI-adjusted) associated with prediabetes using a myriad of definitions, which displayed substantial variability in prevalence (**Figure 3A-B, Supplementary table 4**). First, we observed that IFG —but not high HbA1c— was associated with prevalent arterial hypertension. Moreover, almost all prediabetes definitions displayed roughly comparable estimates for prevalent hypercholesterolemia and hypertriglyceridemia, except for the definition suggested by the IEC, which was poorly associated with these conditions. For insulin resistance, the stronger association was observed with the IFG-WHO definition and the weaker association with the high HbA1c-IEC definition. Prediabetes was also associated with metabolic syndrome, and individuals with alterations in both ADA criteria had a higher likelihood of having this condition. Lastly, we found no association with history of cardiovascular disease for any of the definitions of prediabetes (**Figure 3C**). All odds ratios with 95% confidence intervals are specified in **Supplementary Table 5**.

**FIGURE 3.**
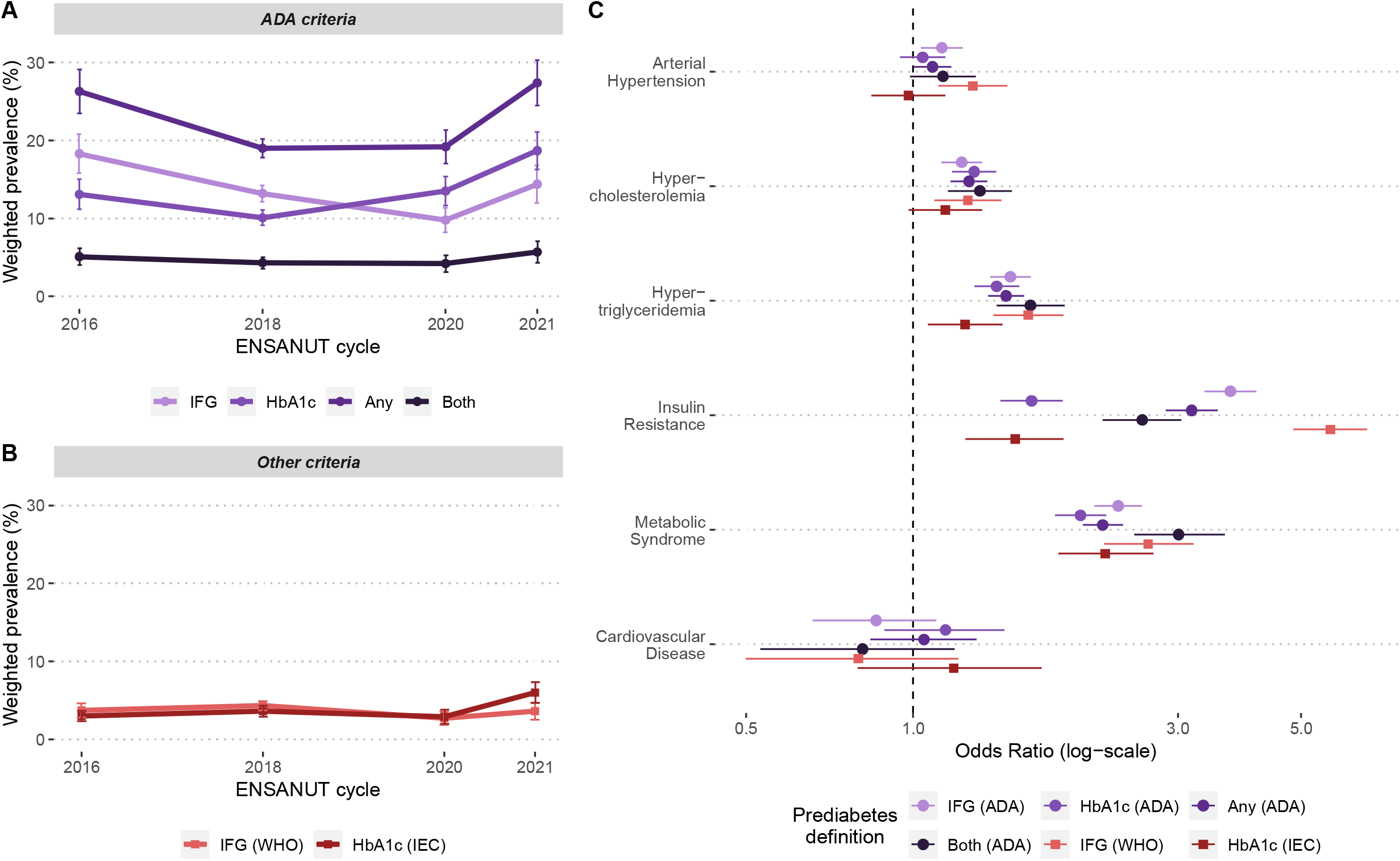
Prevalence of prediabetes in Mexico from 2016 to 2021 according to different definitions used by the ADA (A) and by other organizations (B). We also present an odds ratio plot displaying the association between each of these definitions with multiple cardiovascular outcomes, adjusted for age, sex, and body mass index. <u>Abbreviations</u>. ADA: American Diabetes Association. IEC: SInternational Expert Committee. WHO: World Health Organization. IFG: Impaired fasting glucose.

## DISCUSSION

In this serial cross-sectional study of nationally representative surveys spanning 2016-2021, totaling 21,442 Mexican adults, we found that prediabetes by any definition currently affects 27.3% of the adult population. While the prevalence of prediabetes has decreased over time overall, largely driven by a decrease in IFG using both ADA and WHO criteria, we observed a consistent increase in prevalence using HbA1c, irrespective of the specific criteria. We also observed a consistent increase in prediabetes prevalence among adults over 40 years, individuals self-identified as indigenous, and individuals with central obesity or living in urban areas. Finally, clustering of cardio-metabolic conditions varied widely using different prediabetes definitions, which highlights the need for a consensus to better characterize prediabetes-associated cardiometabolic burden.

### Prevalence of prediabetes

Providing a precise estimate of prediabetes prevalence has been challenging, as estimates widely vary according to definition and study settings. In 2019, the IDF estimated that 7.5% of adults worldwide had IGT, with the highest prevalence (13.8%) observed in North America and the Caribbean (30). Furthermore, using data from the National Health and Nutrition Examination Survey (NHANES) from 2015-2016 (2), prevalence of prediabetes in the U.S. was estimated to be as high as 43% with the ADA-IFG definition (compared to a range of 9.8-18.3% in our study), 15.4% with WHO-IFG (2.7-4.3% in our study), 12.3% with ADA-HbA1c (10.1-18.7% in our study), and 4.3% with IEC-HbA1c (2.9-6.1% in our study). A report from the Center of Disease Control and Prevention (CDC) using data from NHANES 2017-2020 (31) showed that prevalence using either high HbA1c or IFG (ADA-Any) was 38% (19.0-27.3% in our study) and 10.8% using both (4.2-5.7% in our study). These discrepancies could be attributed to differences in study design, sampling, and analysis (e.g., some studies used participants aged ≥18 years and provided an age-adjusted prevalence) or due to ethnic differences, which needs further exploration. Although we did not find a consistent increase in the overall prevalence of prediabetes, we observed an increase using the ADA-HbA1c and IEC-HbA1c definitions and an increase in the prevalence of concordant IFG and high HbA1c over time. Indeed, reports from different regions of the world suggest that the prevalence of prediabetes has increased over the last few decades regardless of the definition; a trend projected to continue(2,30).

### Factors associated to increased prediabetes prevalence

We also found that participants of older age, with obesity, and living in urban areas are more likely to have prediabetes than other populations. This finding is consistent with previous studies that report a strong association of age and obesity with prediabetes (2). Moreover, a similar report from ENSANUT using diabetes as the primary outcome also found urban area and limited access to education to significantly increase its prevalence (32), outlining the importance of sociodemographic and structural factors in the development of glucose metabolism disturbances. Interestingly, we found that Indigenous participants have lower rates of obesity and insulin resistance, which aligns with an earlier study reporting a reduced prevalence of prediabetes and diabetes (defined only with FPG and OGTT) in the Indigenous Mexican population (33). Nonetheless, in our study, Indigenous participants had an elevated prevalence of high HbA1c and were older, had higher triglycerides, and lower HDL-cholesterol. Indigenous participants also showed lower values of HOMA2-IR and HOMA2-β, which could indicate a certain degree of insulin deficiency rather than insulin resistance in this population. Thus, results are not only explained by socioeconomic factors, but probably also by inherent differences in the pathophysiology of glucose metabolism disturbances in Indigenous population, possibly driven by genetic ancestry. These results also give rise to the idea that certain populations (allegedly due to ethnic and racial differences) are at higher risk of specific diabetes phenotypes (34–36), which should be further explored in future studies.

### Cardiometabolic risk conferred by prediabetes

We also evaluated the risk conferred by various definitions and observed that WHO-IFG and the combination of both ADA criteria were associated with the highest risk. Notably, IEC-HbA1c displayed the weaker associations, and no definition was associated with cardiovascular disease. Previously, Tabák, et al. delineated a timeline of the events that tie initial glycemic dysregulation to the onset of diabetes, in which there is a prolonged period of impaired insulin sensitivity and secretion even before the biochemical criteria for prediabetes is met (37). Thus, subjects diagnosed with prediabetes are likely to already present with insulin resistance and β-cell dysfunction, which is why some authors have suggested changes in the diagnostic approach of high-risk subjects to detect earlier disturbances in glucose homeostasis (e.g., replace 2-hour glucose with 1-hour glucose in OGTT) (38). During this prediabetic period, insulin resistance interacts with other risk factors, such as visceral obesity, to precipitate an unfavorable metabolic state characterized by a decline in β-cell mass and function, increased lipolysis, changes in adipokine profiles, systemic inflammation, and endothelial dysfunction, among others (37,39,40). The intersection of these variables is reflected by the association of prediabetes with dyslipidemia, hypertension, and metabolic syndrome in our results. Eventually, a subset of these individuals may progress to overt diabetes —which further accelerates these mechanisms— and may ultimately develop micro and macrovascular complications (37). Conversely, a significant number of individuals with prediabetes may never develop diabetes at all, or even revert to normoglycemia (37), which is often influenced by lifestyle or pharmacological interventions (2). Of note, a recent study by Rooney, et al., found that older adults with prediabetes display low rates of progression to diabetes and high rates of regression to normoglycemia (41), which may generate debate on the utility of the diagnosis of prediabetes in older subjects for risk stratification. Nonetheless, several meta-analyses using data from prospective studies have shown that all prediabetes definitions increase the risk of incident diabetes (42,43), cardiovascular disease, and mortality (44), with WHO-IFG and HbA1c-based definitions showcasing the highest risk in most cases.

In this study, we found no association between prediabetes and cardiovascular disease, however, the cross-sectional design of our study may fail to capture the long-term effects of these abnormalities. Furthermore, this discordance in prevalence and conferred risk driven by different definitions can also be attributed to the inherent properties of each diagnostic test and threshold. For instance, high HbA1c has showed increased specificity for diagnosis of prediabetes and is likely to identify individuals who also meet the other two criteria (2). However, this lowers its utility for screening in the general population, which is why the WHO does not recommend using HbA1c for diagnosis of prediabetes (13). In this regard, IFG is highly sensitive, but there is skepticism in the use of lower cut-offs (i.e., 100-125 mg/dL), as this increases the risk of false positives and possibly captures individuals with lower cardiometabolic risk. Preferably, various diagnostic tests should be used to tailor prevention and therapeutic strategies. For the diagnosis of diabetes mellitus, the ADA recommends using two different tests and not to rely on isolated elevations of a single measurement (21), and it has been questioned whether this recommendation should be extended to prediabetes; our results show that combining multiple criteria (ADA-both) does improve the association with metabolic disease, while reducing the proportion of detected individuals.

### Strengths and limitations

This study is, to our knowledge, the first to estimate prevalence of different prediabetes definitions in the Mexican population using precise and nationally representative data spanning multiple years and considering state-level conglomerations. Given the large burden of diabetes and metabolic diseases in the Mexican population, our estimates are likely to provide useful information for policy makers regarding screening strategies for prediabetes and to identify populations with higher prediabetes prevalence. However, we also acknowledge some limitations which should be considered to adequately interpret our results. First, data on OGTT was not available in ENSANUT; therefore, the overall prevalence of prediabetes may be underestimated as a result. Second, while probabilistic surveys are essential to provide reliable estimates at a national-level, the cross-sectional setting of this study hinders the assessment of long-term risk of prediabetes and its different definitions and further studies are needed to evaluate the longitudinal changes in prediabetes status, as well as the risk of incident diabetes, cardiometabolic disease, and all-cause and cause-specific mortality in individuals with prediabetes using different diagnostic criteria.

## Conclusion

In summary, we observed a high prevalence of prediabetes in the Mexican population which varied widely according to the diagnostic test and cut-off being used. Overall, prevalence of IFG decreased over time, with increases observed in prediabetes as defined by HbA1c criteria. Nevertheless, our results emphasize the importance of early detection and prevention of alterations in glucose metabolism (particularly in populations at high risk), as prediabetes is associated with increased risk of poor cardiometabolic outcomes regardless of the definition. This also highlights the urge to conduct more research and harmonize the definition of prediabetes, considering that the prevalence of this condition will likely increase in the coming years in LMICs.

## Data Availability

All code, datasets and materials are available for reproducibility of results at http://github.com/oyaxbell/prediabetes_ensanut/

## ACKNOWLEDGMENTS

CAFM and JPE are enrolled at the PECEM Program of the Faculty of Medicine at UNAM. CAFM and DRG are supported by CONACyT.

## AUTHOR CONTRIBUTIONS

Research idea and study design: CDPC, CAFM, OYBC; data acquisition and processing: RR, CDPC, CAFM, NEAV, AVV, LFC, DRG; statistical analysis: CDPC, CAFM, OYBC; analysis/interpretation: CDPC, CAFM, OYBC, NEAV, AVV, LFC, DRG; manuscript drafting: JPE, MRBA, PSC, ANL, KCH, LCQ, CDPC, CAFM, JAS, OYBC; supervision or mentorship: JAS, OYBC. Each author contributed important intellectual content during manuscript drafting or revision and accepts accountability for the overall work by ensuring that questions pertaining to the accuracy or integrity of any portion of the work are appropriately investigated and resolved.

