## Supplementary Material for "Prevalence of untreated prediabetes and glucose metabolism disturbances in Mexico: An analysis of nationally representative surveys spanning 2016-2021"

**SUPPLEMENTARY METHODS**

**National Health and Nutrition Survey**

The Mexican National Health and Nutrition Survey (ENSANUT) comprises a series of population-based surveys that provide precise information on the health and nutritional status of Mexican population, as well as on the performance of our health care system. ENSANUT uses a two-stage probabilistic cluster stratified sampling based on households and individuals from each group of interest (adults [≥20 years], adolescents [10-19 years], school age [5-9 years], and preschool age [0-4 years]), and it is representative at a regional (North, Center, Mexico City, and South), and national level for rural and urban areas. ENSANUT has been conducted in 2006, 2012, 2016, 2018, 2020, 2021, and 2022. Sociodemographic and dwelling data is obtained from each household, and individual participants undergo a standardized health questionnaire that is applied in face-to-face interviews, followed by a physical exam including measurement of blood pressure and anthropometry (height in centimeters, weight in kilograms, and waist circumference in centimeters [not available for 2020]) by trained personnel. The physical exam was performed only in a subset of participants for the 2018 (40%) and 2021 (80 %) surveys. Additionally, a random subset of each cycle was selected for venous blood collection for biochemical evaluation of the following serum laboratory parameters: albumin, creatinine, uric acid, triglycerides, total cholesterol, LDL-cholesterol, HDL-cholesterol, glucose, insulin, glycated hemoglobin, and from 2020 onwards SARS-CoV-2 serology. All questionnaires and procedures were reviewed and approved by the Ethics, Investigation and Biosecurity Committee from the National Institute of Public Health for each year of survey (1–4).

**Social Lag Index (SLI) and Density-Independent Social Lag Index (DISLI)**

We used data from the National Council for Evaluation of Social Development Policy (CONEVAL), which provides state-level estimates of the 2020 social lag index (SLI), which is a composite measure of access to education, health care, dwelling quality, and basic services in Mexico (5). To assess marginalization independently of urbanization, we extracted mean urban population density from CONEVAL and regressed it onto SLI and obtained the residuals which represent a density-independent social lag index (DISLI) (6,7). We then categorized states into DISLI quartiles ("Very-Low", "Low", "Middle", and "High") and the prevalence of prediabetes among high DISLI states was contrasted against the rest of states.

**SUPPLEMENTARY TABLES**

**SUPPLEMENTARY TABLE 1.** Population characteristics for each year of survey. Data is presented as median (with interquartile range) or as absolute frequency (with percentage). Abbreviations. BMI: Body mass index. WC: Waist circumference. FPG: Fasting plasma glucose. HOMA2-IR: Homeostatic model assessment for insulin resistance.

| **Parameter** | **ENSANUT 2016 N = 8295** | **ENSANUT 2018 N = 42565** | **ENSANUT 2020 N = 2373** | **ENSANUT 2021 N = 13402** |
| --- | --- | --- | --- | --- |
| **Sociodemographic** | | | | |
| Age (years) | 44 (33–58) | 43 (32–57) | 44 (32–58) | 44 (32–57) |
| Sex (women) | 5437 (65.7%) | 23300 (54.7%) | 1410 (59.4%) | 8166 (60.9%) |
| Never smokers | 4253 (51.5%) | 26114 (61.5%) | – | 8914 (66.5%) |
| Indigenous language | 964 (11.6%) | 3021 (7.1%) | 125 (5.3%) | 626 (4.7%) |
| Urban area | 4155 (50.1%) | 30979 (72.8%) | 1812 (76.4%) | 10216 (76.2%) |
| **Physical exam and laboratory data** | | | | |
| BMI (kg/m^2) | 28 (24.8–31.7) | 28.4 (25.1–32.1) | 28.3 (25.1–32.2) | 28.7 (25.2–32.6) |
| WC (cm) | 94.2 (86.3–102.8) | 96 (87.6–104.7) | – | 97 (88.3–106.2) |
| FPG (mg/dL) | 95 (88–105) | 91 (83–103) | 89 (82–100) | 93.15 (86.3–103.5) |
| HbA1c (%) | 5.4 (5.1–5.7) | 5.3 (5–5.7) | 5.4 (5.1–5.7) | 5.5 (5.2–5.9) |
| HOMA2-IR | 1.1 (0.7–1.7) | 1.1 (0.7–1.9) | 1.1 (0.7–1.9) | 1.1 (0.7–1.8) |
| **Comorbidities** | | | | |
| Hypertension | 2241 (27%) | 12042 (28.3%) | 420 (18.8%) | 3245 (24.2%) |
| Hyper-cholesterolemia | 1279 (34.8%) | 4243 (33.5%) | 637 (27.8%) | 539 (26.2%) |
| Hyper-triglyceridemia | 2047 (55.7%) | 7324 (57.9%) | 1172 (51.2%) | 1014 (49.3%) |
| Cardiovascular disease | 249 (3%) | 1055 (2.5%) | – | 255 (1.9%) |

**SUPPLEMENTARY TABLE 2.** Prevalence rate ratios (RR) with 95% confidence intervals from mixed-effects Poisson models using ***high HbA1c as defined by the ADA*** as the diagnostic criteria for prediabetes. We included a random slope for state of origin to address state-level conglomeration. Abbreviations. BMI: Body mass index. DISLI: Density-independent social lag index.

| **Model** | **Predictor** | **RR (95% CI)** |
| --- | --- | --- |
| **Age group**  **(years)** | 20-39 | Ref |
|  | 40-59 | **3.152 (2.595 - 3.829)** |
|  | ≥60 | **4.322 (3.517 - 5.313)** |
|  | ENSANUT year | 1.002 (0.941 - 1.067) |
|  | Year*Age 40-59 | 1.048 (0.975 - 1.128) |
|  | Year*Age ≥60 | 1.061 (0.983 - 1.146) |
| **Sex** | Men | Ref |
|  | Women | 1.087 (0.937 - 1.260) |
|  | ENSANUT year | 1.032 (0.987 - 1.079) |
|  | Year*Women | 1.025 (0.971 - 1.082) |
| **BMI categories** | Normal weight | Ref |
|  | Overweight | **1.353 (1.089 - 1.680)** |
|  | Obesity | **1.955 (1.590 - 2.405)** |
|  | ENSANUT year | 0.936 (0.870 - 1.007) |
|  | Year*Overweight | **1.098 (1.007 - 1.196)** |
|  | Year*Obesity | **1.153 (1.063 - 1.250)** |
| **Waist circumference** | Normal | Ref |
|  | Central obesity | **1.664 (1.319 - 2.100)** |
|  | ENSANUT year | **0.876 (0.787 - 0.975)** |
|  | Year*Obesity | **1.221 (1.093 - 1.364)** |
| **Smoking status** | Never smoker | Ref |
|  | Former smoker | **1.239 (1.057 - 1.452)** |
|  | Current smoker | **0.791 (0.630 - 0.993)** |
|  | ENSANUT year | **1.083 (1.044 - 1.122)** |
|  | Year*Former | **0.911 (0.850 - 0.976)** |
|  | Year*Current | 1.030 (0.946 - 1.121) |
| **Indigenous language** | No | Ref |
|  | Yes | **1.369 (1.090 - 1.696)** |
|  | ENSANUT year | 1.058 (1.039 - 1.088) |
|  | Year*Yes | 0.920 (0.835 - 1.014) |
| **DISLI** | Low-Middle | Ref |
|  | High | **1.170 (1.004 - 1.364)** |
|  | ENSANUT year | **1.073 (1.041 - 1.106)** |
|  | Year*High | **0.939 (0.885 - 0.997)** |
| **Area** | Rural | Ref |
|  | Urban | 0.912 (0.800 - 1.040) |
|  | ENSANUT year | 0.959 (0.920 - 1.000) |
|  | Year*Urban | **1.091 (1.050 - 1.132)** |

**SUPPLEMENTARY TABLE 3.** Prevalence rate ratios (RR) with 95% confidence intervals from mixed-effects Poisson models using ***impaired fasting glucose as defined by the ADA*** as the diagnostic criteria for prediabetes. We included a random slope for state of origin to address state-level conglomeration. Abbreviations. BMI: Body mass index. DISLI: Density-independent social lag index.

| **Model** | **Predictor** | **RR (95% CI)** |
| --- | --- | --- |
| **Age group**  **(years)** | 20-39 | Ref |
|  | 40-59 | **1.966 (1.705 - 2.266)** |
|  | ≥60 | **1.941 (1.641 - 2.295)** |
|  | ENSANUT year | **0.872 (0.832 - 0.915)** |
|  | Year*Age 40-59 | 1.014 (0.955 - 1.077) |
|  | Year*Age ≥60 | 1.020 (0.951 - 1.094) |
| **Sex** | Men | Ref |
|  | Women | 0.999 (0.881 - 1.132) |
|  | ENSANUT year | **0.900 (0.863 - 0.938)** |
|  | Year*Women | 0.970 (0.921 - 1.022) |
| **BMI categories** | Normal weight | Ref |
|  | Overweight | **1.876 (1.543 - 2.281)** |
|  | Obesity | **2.733 (2.265 - 3.298)** |
|  | ENSANUT year | **0.880 (0.819 - 0.946)** |
|  | Year*Overweight | 0.976 (0.897 - 1.062) |
|  | Year*Obesity | 1.012 (0.934 - 1.097) |
| **Waist circumference** | Normal | Ref |
|  | Central obesity | **2.174 (1.765 - 2.678)** |
|  | ENSANUT year | **0.868 (0.788 - 0.957)** |
|  | Year*Obesity | 1.035 (0.935 - 1.145) |
| **Smoking status** | Never smoker | Ref |
|  | Former smoker | 0.999 (0.870 - 1.147) |
|  | Current smoker | 0.819 (0.668 - 1.005) |
|  | ENSANUT year | **0.892 (0.860 - 0.925)** |
|  | Year*Former | 1.028 (0.962 - 1.099) |
|  | Year*Current | 1.010 (0.923 - 1.105) |
| **Indigenous language** | No | Ref |
|  | Yes | 1.187 (0.974 - 1.447) |
|  | ENSANUT year | **0.893 (0.869 - 0.917)** |
|  | Year*Yes | **0.844 (0.761 - 0.936)** |
| **DISLI** | Low-Middle | Ref |
|  | High | 0.980 (0.861 - 1.117) |
|  | ENSANUT year | **0.881 (0.855 - 0.909)** |
|  | Year*High | 1.026 (0.971 - 1.085) |
| **Area** | Rural | Ref |
|  | Urban | 1.009 (0.901 - 1.131) |
|  | ENSANUT year | **0.841 (0.808 - 0.876)** |
|  | Year*Urban | **1.047 (1.009 - 1.087)** |

**SUPPLEMENTARY TABLE 4.** Annual prevalence with 95% confidence intervals for prediabetes as defined by the ADA, WHO and IEC. Abbreviations. ADA: American Diabetes Association. IEC: International Expert Committee. WHO: World Health Organization. IFG: Impaired fasting glucose.

| **Definition** | **Year** | **Prevalence (%)** | **95% CI** |
| --- | --- | --- | --- |
| **ADA-IFG** | 2016 | 18.3 | 15.8 – 20.8 |
|  | 2018 | 13.2 | 12.1 – 14.2 |
|  | 2020 | 9.8 | 8.2 – 11.4 |
|  | 2021 | 14.4 | 12.0 – 16.8 |
| **ADA-A1c** | 2016 | 13.1 | 11.2 – 15.0 |
|  | 2018 | 10.1 | 9.1 – 11.1 |
|  | 2020 | 13.5 | 11.6 – 15.4 |
|  | 2021 | 18.7 | 16.3 – 21.1 |
| **ADA-Any** | 2016 | 26.3 | 23.5 – 29.1 |
|  | 2018 | 19.0 | 17.8 – 20.2 |
|  | 2020 | 19.2 | 17.0 – 21.3 |
|  | 2021 | 27.3 | 24.4 – 30.2 |
| **ADA-Both** | 2016 | 5.1 | 4.0 – 6.2 |
|  | 2018 | 4.3 | 3.5 – 5.0 |
|  | 2020 | 4.2. | 3.1 – 5.3 |
|  | 2021 | 5.7 | 4.3 – 7.1 |
| **WHO-IFG** | 2016 | 3.7 | 2.8 – 4.6 |
|  | 2018 | 4.3 | 3.7 – 4.9 |
|  | 2020 | 2.7 | 1.9 – 3.6 |
|  | 2021 | 3.6 | 2.5 – 4.7 |
| **IEC-A1c** | 2016 | 3.0 | 2.3 – 3.7 |
|  | 2018 | 3.6 | 2.9 – 4.3 |
|  | 2020 | 2.9 | 2.0 – 3.8 |
|  | 2021 | 6.1 | 4.8 – 7.4 |

**SUPPLEMENTARY TABLE 5.** Odds ratios (OR) with 95% confidence intervals from logistic regression models to predict cardiometabolic outcomes using diverse prediabetes definitions (estimates for prediabetes by any ADA criteria and diabetes are shown in bold for comparisons). All models were adjusted for age, sex, and body mass index. Abbreviations. ADA: American Diabetes Association. IEC: International Expert Committee. WHO: World Health Organization. IFG: Impaired fasting glucose.

| **Outcome** | **Predictor** | **OR (95% CI)** |
| --- | --- | --- |
| **Hypertension** | IFG (ADA) | 1.127 (1.033 – 1.230) |
|  | HA1c (ADA) | 1.041 (0.948 – 1.143) |
|  | ***Any (ADA)*** | ***1.084 (1.003 – 1.172)*** |
|  | Both (ADA) | 1.133 (0.988 – 1.298) |
|  | IFG (WHO) | 1.282 (1.110 – 1.479) |
|  | HbA1c (IEC) | 0.981 (0.842 – 1.144) |
|  | ***Diabetes mellitus*** | ***1.865 (1.730 – 2.010)*** |
| **Hypercholesterolemia** | IFG (ADA) | 1.223 (1.124 – 1.331) |
|  | HbA1c (ADA) | 1.289 (1.175 – 1.414) |
|  | ***Any (ADA)*** | ***1.262 (1.169 – 1.362)*** |
|  | Both (ADA) | 1.320 (1.155 – 1.508) |
|  | IFG (WHO) | 1.256 (1.091 – 1.444) |
|  | HbA1c (IEC) | 1.144 (0.981 – 1.332) |
|  | ***Diabetes mellitus*** | ***1.387 (1.276 – 1.507)*** |
| **Hypertriglyceridemia** | IFG (ADA) | 1.498 (1.377 – 1.629) |
|  | HbA1c (ADA) | 1.415 (1.289 – 1.553) |
|  | ***Any (ADA)*** | ***1.471 (1.365 – 1.585)*** |
|  | Both (ADA) | 1.629 (1.416 – 1.877) |
|  | IFG (WHO) | 1.613 (1.396 – 1.867) |
|  | HbA1c (IEC) | 1.241 (1.064 – 1.450) |
|  | ***Diabetes mellitus*** | ***2.407 (2.205 – 2.628)*** |
| **Insulin Resistance** | IFG (ADA) | 3.731 (3.347 – 4.158) |
|  | HbA1c (ADA) | 1.636 (1.436 – 1.860) |
|  | ***Any (ADA)*** | ***3.176 (2.851 – 3.539)*** |
|  | Both (ADA) | 2.588 (2.195 – 3.044) |
|  | IFG (WHO) | 5.650 (4.844 – 6.585) |
|  | HbA1c (IEC) | 1.528 (1.243 – 1.868) |
|  | Diabetes mellitus | ***–*** |
| **Metabolic Syndrome** | IFG (ADA) | 2.341 (2.123 – 2.585) |
|  | HbA1c (ADA) | 2.004 (1.802 – 2.231) |
|  | ***Any (ADA)*** | ***2.196 (2.022 – 2.387)*** |
|  | Both (ADA) | 3.009 (2.503 – 3.646) |
|  | IFG (WHO) | 2.652 (2.211 – 3.204) |
|  | HbA1c (IEC) | 2.218 (1.828 – 2.713) |
|  | Diabetes mellitus | ***–*** |
| **Cardiovascular disease** | IFG (ADA) | 0.858 (0.660 – 1.103) |
|  | HbA1c (ADA) | 1.145 (0.888 – 1.462) |
|  | ***Any (ADA)*** | ***1.047 (0.838 – 1.302)*** |
|  | Both (ADA) | 0.811 (0.531 – 1.189) |
|  | IFG (WHO) | 0.798 (0.500 – 1.207) |
|  | HbA1c (IEC) | 1.184 (0.794 – 1.705) |
|  | ***Diabetes mellitus*** | ***1.379 (1.139 – 1.670)*** |

**SUPPLEMENTARY FIGURES**

**Supplementary Figure 1.** Flowchart of participant selection for each year of survey. We display the total number of participants surveyed per year (first row), and the number of them which completed the health questionnaire (second row). The ***general population*** of our study is therefore comprised of participants ≥20 years old that completed the health questionnaire (third row), while the ***laboratory subset*** includes those with complete weights for venous blood and sampling stratum (fourth row). This diagram was designed using resources created by *Freepik* from [www.flaticon.com](http://www.flaticon.com)

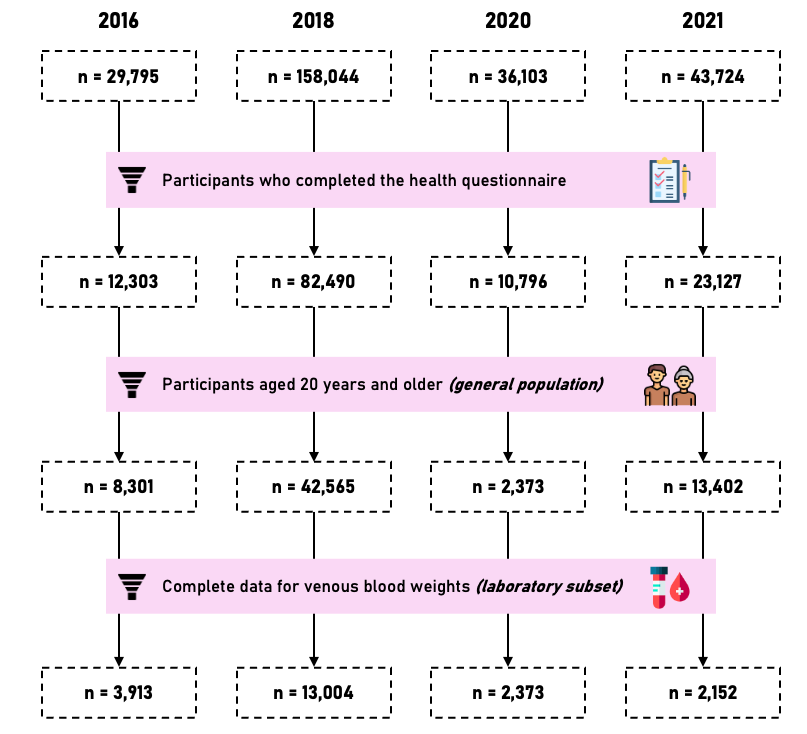

**Supplementary Figure 2.** Percentage of missing values in the general population (A) and in the laboratory subset (B) for all variables included in our study for each year of survey. Abbreviations. BMI: Body mass index. WC: Waist circumference. SBP: Systolic blood pressure. DBP: Diastolic blood pressure. HX: History (previous medical diagnosis) of the disease. CVD: Cardiovascular disease. HDL-cholesterol: High density lipoprotein cholesterol.

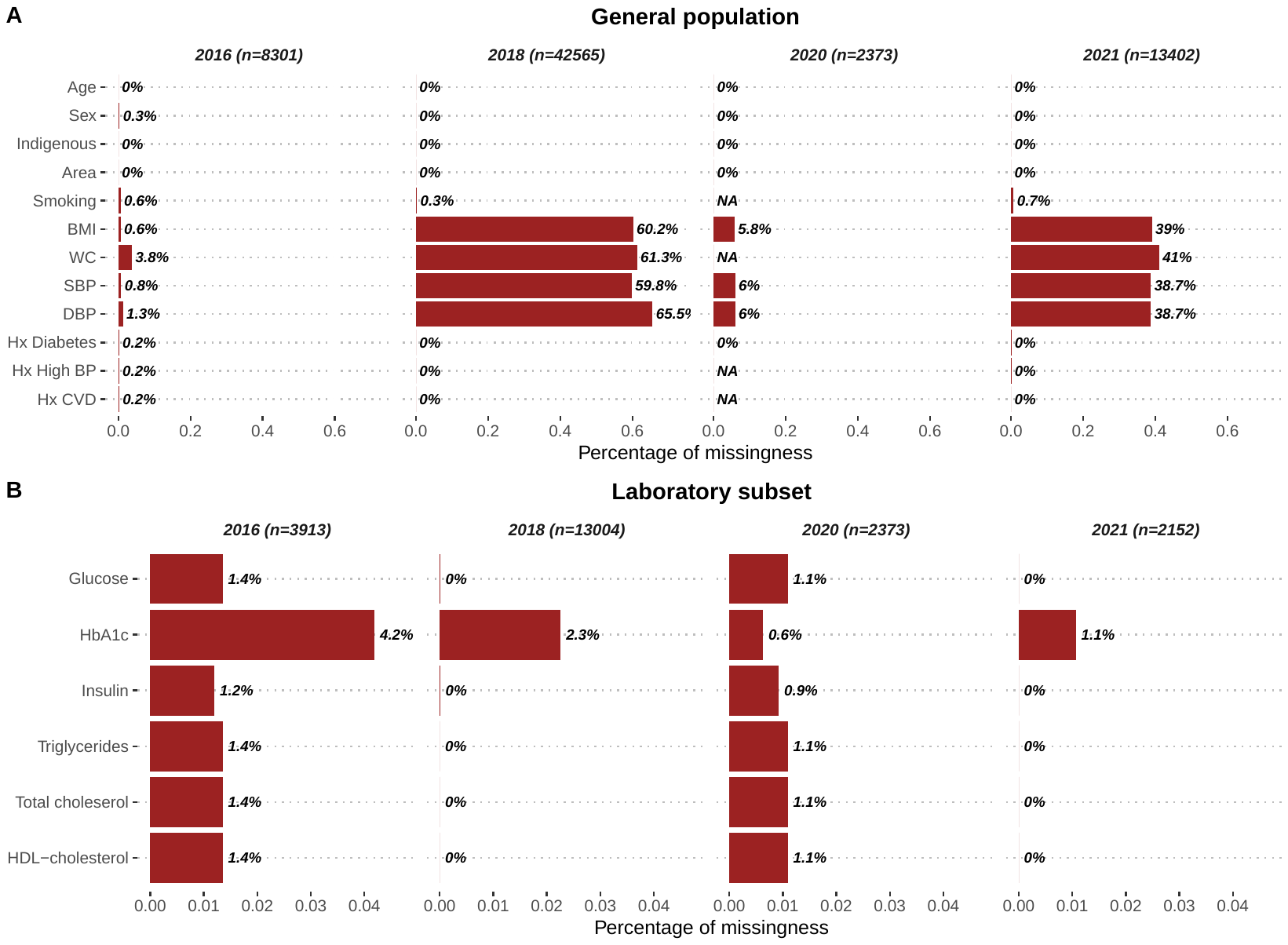

**Supplementary Figure 3.** Change in prevalence of prediabetes (high HbA1c and IFG criteria) from 2016 to 2021 in Mexico stratified by multiple modifying factors. Abbreviations. BMI: Body mass index. DISLI: Density-independent social lag index. IFG: Impaired fasting glucose.

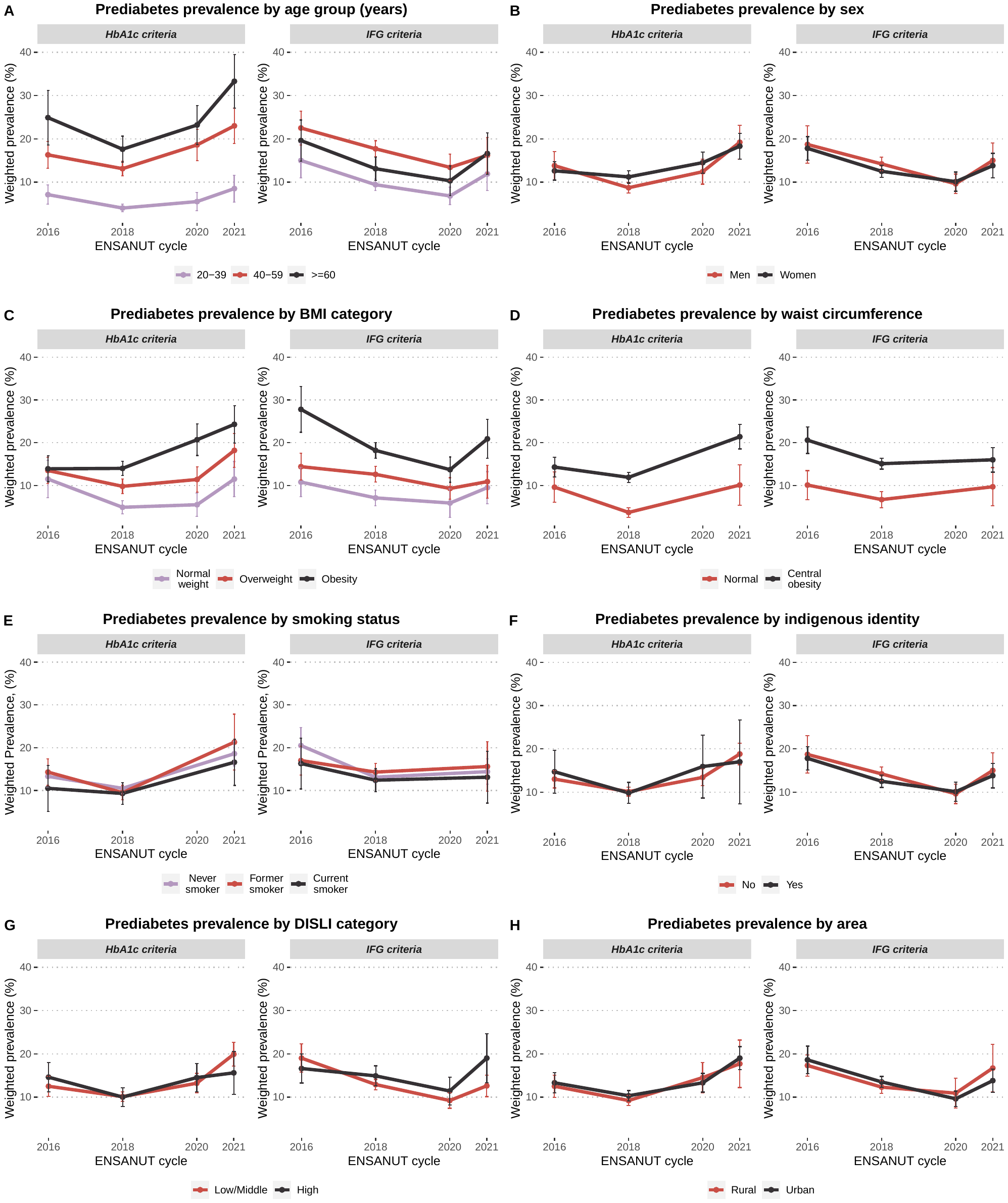

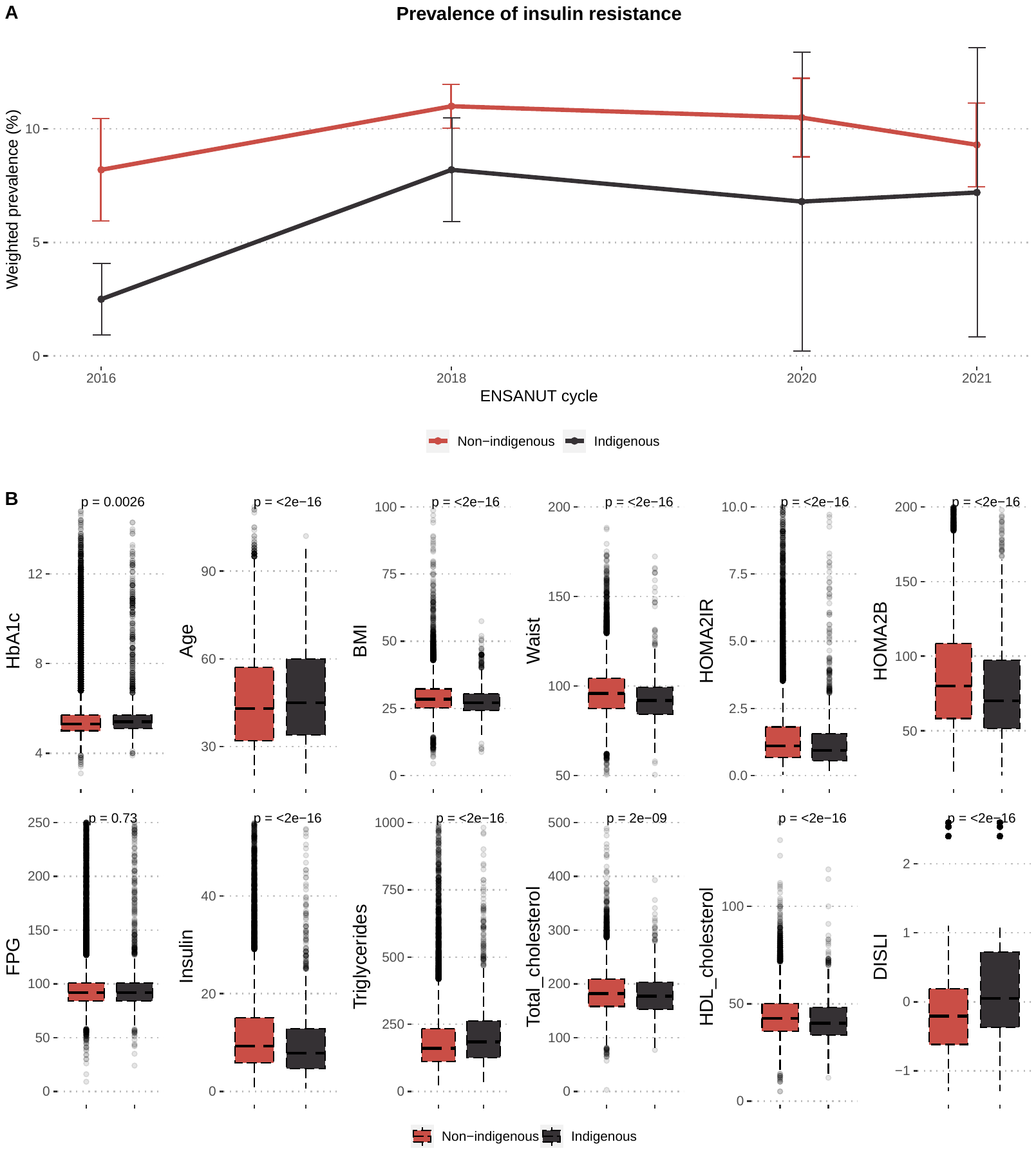
**Supplementary Figure 4.** Prevalence of insulin resistance (defined as HOMA2-IR >2.5) in indigenous (n=1826) and non-indigenous (n=19085) participants from 2016 to 2021 (A). Indigenous individuals display significant differences (Wilcoxon test) in numerous demographic and health-related variables compared to non-indigenous (B). Abbreviations. DISLI: Density-independent social lag index. BMI: Body mass index. SBP: Systolic blood pressure. DBP: Diastolic blood pressure. FPG: Fasting plasma glucose.
